# GWAS for Periodontitis Phenotypes Using Multi-Ancestry All of Us Research Platform

**DOI:** 10.1101/2025.09.07.25335133

**Authors:** Keith Sanders, Ardalan Naseri, Chun-Teh Lee, Junichi Iwata, Yixuan He, Bunmi Tokede, Muhammad Walji, Degui Zhi, Laila Rasmy

## Abstract

Periodontitis is a multifactorial inflammatory disease whose pathogenesis is associated with intricate interactions between genetic and environmental factors. Leveraging electronic health records data from the All of Us Research Program, we stratified periodontitis by clinically relevant dimensions: stage, grade, and extent. Based on these phenotypes, we performed a multi-ancestry genome-wide association study, focusing on predominant ancestry populations of African, European, and Admixed American. Our study cohort comprised 3,881 periodontitis patients and a control group of 10,760 patients with dental caries and without periodontitis. Ancestry-specific GWAS revealed significant genetic associations (P<5x10^-8^) in periodontitis grade phenotypes at the LINC00294 and CLMN loci in the African ancestry population and also confirmed via the multi-ancestry meta-analysis. In addition, the XYLT1 locus emerged as a significant signal associated with periodontitis grade phenotype in the admixed American GWAS. Our GWAS comparing periodontitis to dental caries in the admixed American population identified several significant loci, including RABGAP1L, previously linked to immune regulation, DCHS2, a cadherin-related gene involved in bone mineralization and tissue morphogenesis, and OSTM1, known to be crucial for bone remodeling. The findings of our study highlight the potential of integrating EHR and genomic data from large-scale biobanks to achieve informative dental phenotyping, uncover novel molecular insights into periodontal disease, and personalize treatment approaches.

## Introduction

Periodontitis is a multifactorial disease characterized by chronic inflammation of the gingival tissue, which destroys the periodontal ligament, alveolar bone, and supporting connective structures of the teeth [1]. The National Health and Nutrition Examination Survey (NHANES) conducted from 2009 to 2014 estimated that approximately 42% of U.S. adults aged 30 and older have periodontitis [2].

Periodontitis has also been associated with an increased risk for systemic diseases, including diabetes and cardiovascular disease [3]. As a complex disease, the pathogenesis of periodontitis is associated with several genetic, environmental, and lifestyle factors [4]. Moreover, the heterogeneous nature of periodontitis leads to varied clinical manifestations, complicating diagnosis and therapeutic strategies [5]. This variability also reflects the disease’s complex interplay with both host-related and environmental factors [6]. To capture its clinical heterogeneity, periodontitis is assessed along three phenotypic dimensions: stage, grade, and extent, reflecting the disease severity, progression rate, and distribution of affected sites following the 2017 World Workshop on the Classification of Periodontal and Peri-Implant Diseases and Conditions (WWC) [7]. Incorporating these phenotypic dimensions into clinical assessment is critical for accurately characterizing disease burden and tailoring personalized treatment plans [8,9].

A key approach to understanding the genetic components of complex diseases has been facilitated by genome-wide association (GWAS) studies [10]. Although prior GWAS have identified over 65 loci associated with periodontitis for ascribing the heritable component of the disease, these efforts were primarily based on relatively small, homogeneous cohorts, limiting their relevance to diverse populations [11]. Moreover, few of these studies have incorporated clinical stratifications, hindering our understanding of how genetic variation influences the heterogeneity of disease presentation.

To alleviate this gap, electronic health record (EHR)-linked biobanks have emerged as powerful platforms for large-scale genetic discovery across diverse, clinically defined populations [12]. These resources enable the analysis of distinct disease characterization at the molecular level and facilitate the identification of novel genetic contributors to complex diseases [13]. Recent genetic studies leveraging large biobank cohorts have advanced periodontitis research, notably identifying four novel loci associated with periodontitis using UK Biobank (UKB) data [14]. Similarly, the GLIDE consortium and UKB identified 47 novel genetic risk loci for dental caries and provided insights into shared genetic pathways with systemic diseases [15]. However, this study captured the broader landscape of dental caries and denture traits. Their periodontitis-specific analysis yielded only one locus (*SIGLEC5*), underscoring the difficulty of detecting robust signals for this condition. These studies exemplify the utility of biobank-scale resources that can successfully identify clinically relevant genetic contributors to oral diseases. The All of Us Research Program (AOURP), launched by the National Institutes of Health (NIH) in 2018 [16], represents a comprehensive biobank resource that has not previously been applied to periodontitis genetics. The AOURP, a precision medicine initiative, aimed to construct a diverse biomedical database in the U.S.

This study leverages the AOURP cohort, which has been standardized to the Observational Medical Outcomes Partnership (OMOP) common data model, as developed by the Observational Health Data Sciences and Informatics (OHDSI) community [17], to conduct ancestry-specific and multi-ancestry genome-wide association analyses across periodontitis stage, grade, and extent. Applying these clinical stratifications directly addresses prior phenotype resolution and cohort diversity limitations. We aim to identify ancestry-informed genetic risk loci that underlie disease heterogeneity. The findings of our study offer a foundation for subsequent research striving to enhance precision phenotyping and to inform the design of targeted prevention and treatment interventions.

## Results

### Study Cohort

Within our primary periodontitis cohort of 3,881 individuals, we derived three distinct phenotypic subgroups stratified by stage, grade, and extent. These groups consisted of 946, 3,816, and 1,723 participants, respectively. The dental caries (control group) participants comprised 10,760 patients exclusively. Participants were predominantly EUR, followed by AFR and AMR. The mean age of 53 was observed in the periodontitis cohort. The mean age of phenotypic subgroups was observed at age 60 in stage cohorts, 53 in grade, and 57 in extent. The mean age for dental caries cohorts skewed slightly younger, calculated as 49. Sex distribution was balanced in periodontitis and dental caries patients at 48% and 44% male, respectively. After phenotypic characteristics delineated subgroups, we observed a heavier skew towards the male sex. A detailed summary of participant numbers per ancestry and cohort across the four GWAS groups is provided in Table 1.

**Table 1.**
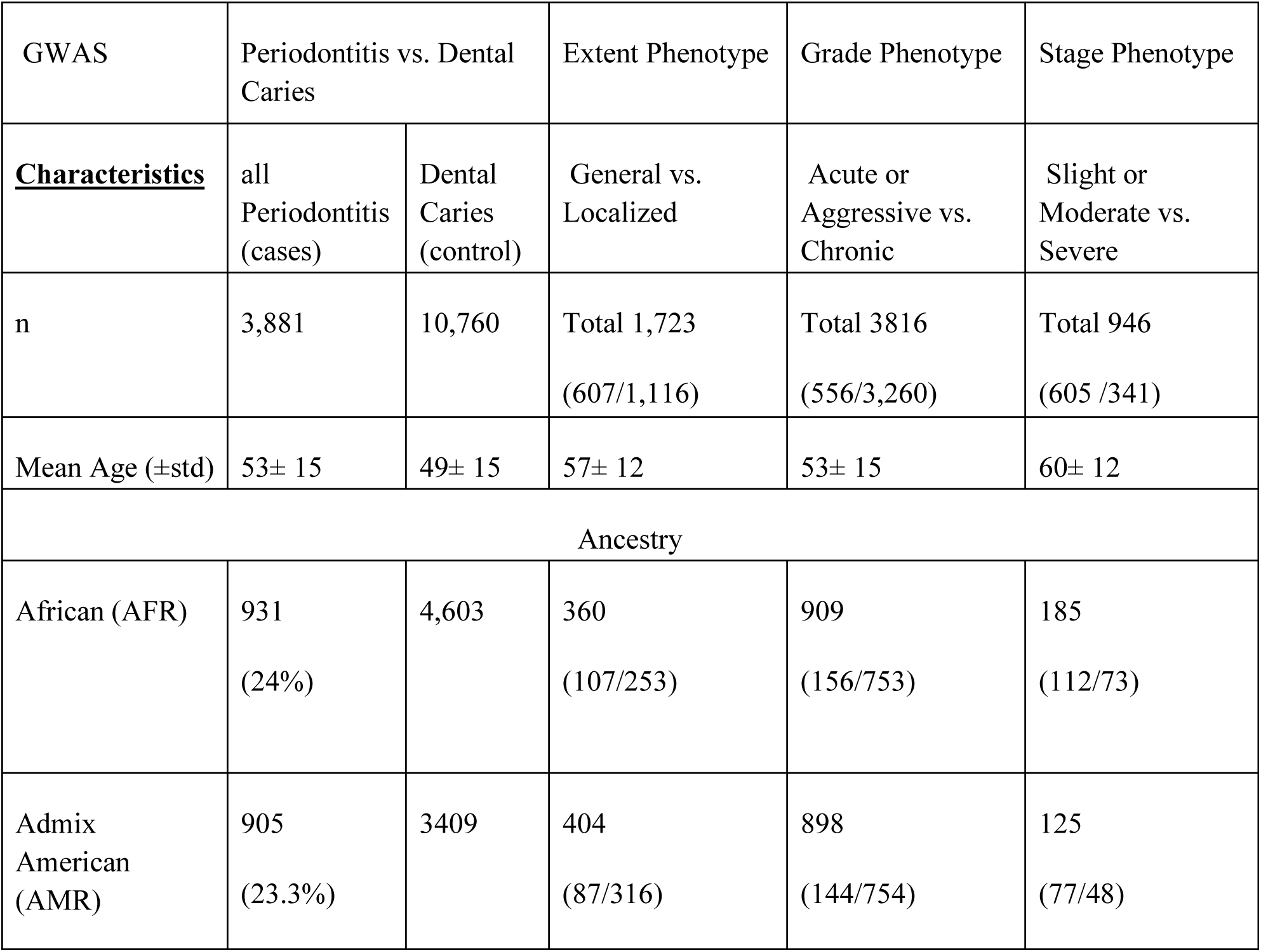
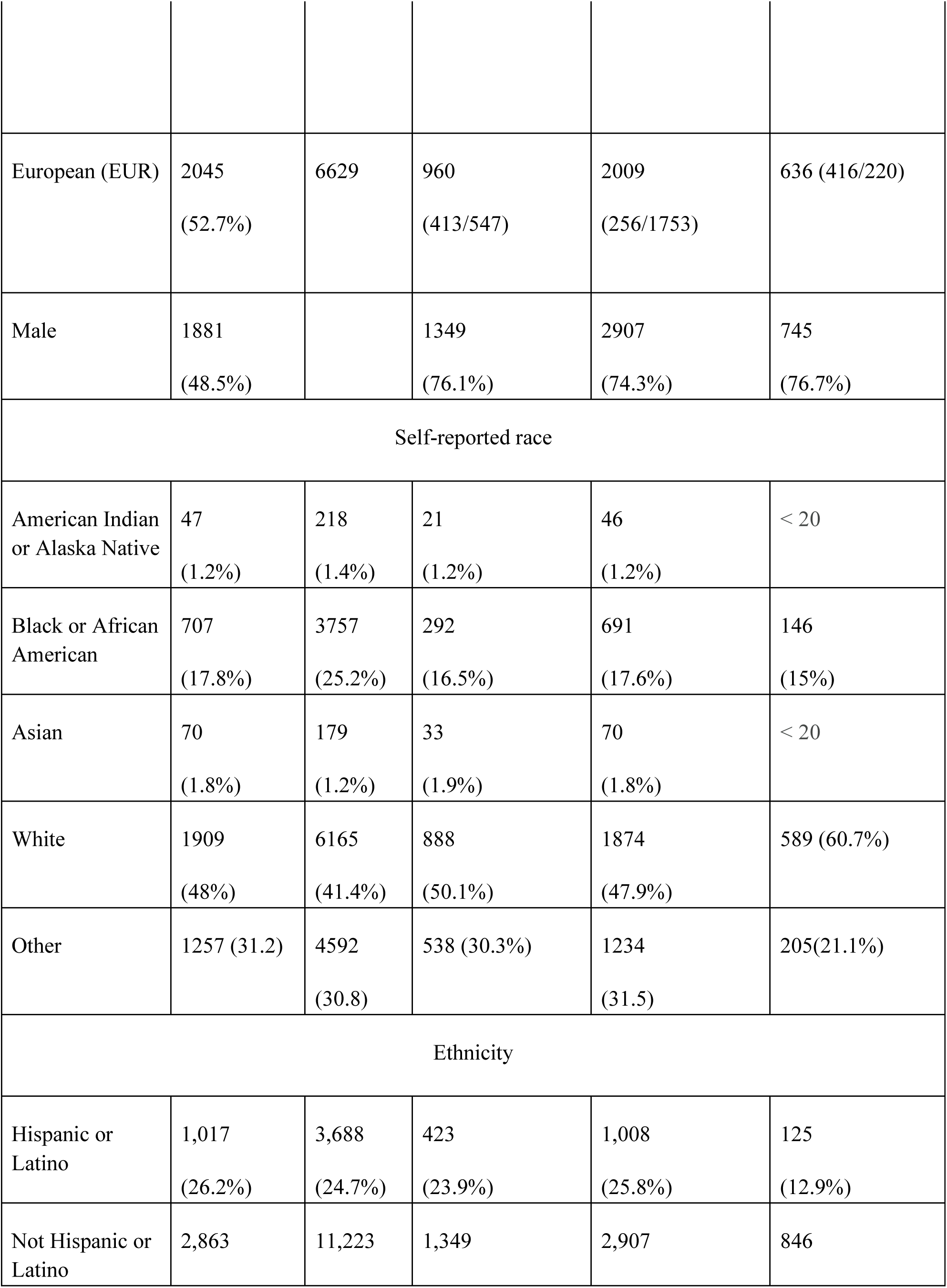
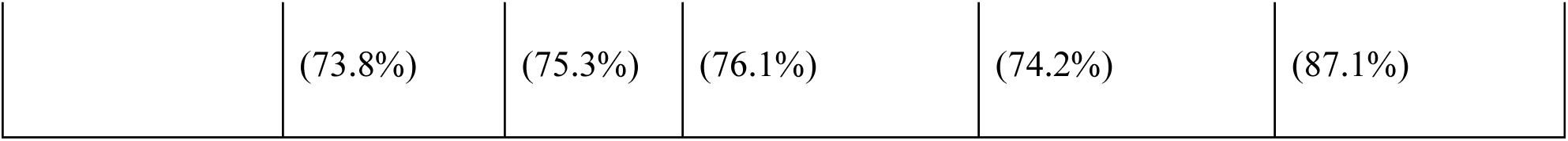
Patient demographic and ancestry distribution across periodontitis and dental caries GWAS phenotypes.

### Genetic Data Quality Assessment

Kinship filtering was applied using the AOURP provided kinship coefficients (kinship >0.1) to remove related participants within each GWAS. This filtering resulted in the exclusion of samples for the periodontitis vs. dental caries GWAS (1,442), for the extent GWAS (N = 335), for the grade GWAS (N = 324), and for the stage phenotype GWAS (N = 32). The final cohort totals for each analysis after these exclusions are presented in Table 1. QQ plots were generated for AFR, AMR, and EUR ancestries, and the multi-ancestry meta-analysis was performed for each GWAS. For the GWAS comparing periodontitis to dental caries (Supplementary Fig. 1), AFR (λ = 0.923) and AMR (λ = 0.907) distributions closely followed expectation, with deviation at the tail consistent with true associations. The EUR analysis (λ = 0.323) produced a muted signal distribution, consistent with fewer observed loci. The meta-analysis (λ = 0.736) demonstrated overall consistency with elevation at the tail for genome-wide loci. For the periodontitis grade phenotype, QQ plots were generated for AFR, AMR, and EUR ancestries and meta-analysis (can be observed in Supplementary Fig. 2. In the AFR analysis (λ = 0.989), the distribution remained consistent with expectation, with deviation at the tail indicating association signals. The AMR analysis (λ = 1.08) showed mild inflation with upward deviation at the furthest tail, consistent with true associations. The distribution was conservative in the EUR analysis (λ = 0.614). The meta-analysis (λ = 0.913) tracked near expectation overall, with tail deviation corresponding to genome-wide loci. For the periodontitis extent phenotype, Extent phenotype QQ plots are found in Supplementary Fig. 3. In the AFR analysis (λ = 0.904), results aligned with the null expectation, with modest tail deviation indicating association signals. The AMR analysis (λ = 0.989) shows a modest increase at the tail. The EUR analysis (λ = 0.296) showed an underpowered distribution, likely driven by fewer detectable signals. The extent phenotype (λ = 0.612) meta-analysis tracked near expectation overall, with deviation at the tail indicating genome-wide associations. For the periodontitis stage phenotype, QQ plots were generated for each ancestry, and the meta-analysis is located in Supplementary Fig. 4. The AFR analysis (λ = 0.911) showed agreement with the null model overall, with modest deviation at the tail. In the AMR analysis (λ = 0.941), results were similar, with a slight increase at the tail consistent with association signals. In the EUR analysis (λ = 0.438), the test statistics appeared deflated, consistent with a smaller sample size and reduced detection of associations. The meta-analysis of the stage phenotype (λ = 0.630) demonstrated consistency with the null, with upward deviation at the tail reflecting genome-wide associations.

### GWAS Findings

Based on GWAS analysis, we identified three loci from the AMR ancestry in all periodontitis cases versus dental caries controls. These genes were ***RABGAP1L*** (rs1183875194, p value = 9.303e-09), ***DCHS2*** (rs10677099, p value = 5.889e-09), and ***OSTM1*** (rs113597243, p value = 3.244e-09). For the periodontitis phenotypes GWAS, we identified three genome-wide significant loci associated with the grade phenotype (Figure 1). In the AFR ancestry, we identified ***LINC00294*** (rs73482834, p value = 3.988e-09) and ***CLMN*** (rs139217059, p value = 1.764e-08). *LINC00294* encodes a long non-coding RNA whose mechanistic role remains under investigation; however, its dysregulation has been linked to altered cell proliferation and apoptosis [18]. *CLMN* encodes calmin, an actin-binding protein, and prior studies have suggested its role in cell proliferation. The AMR ancestry identified ***XTLY1* (**rs147819514, p-value = 3.800e-09) as the nearest gene to the locus. We summarized the ancestry-stratified GWAS findings, including summary statistics and variant annotations for each SNP in Table 2.

**Figure 1.**
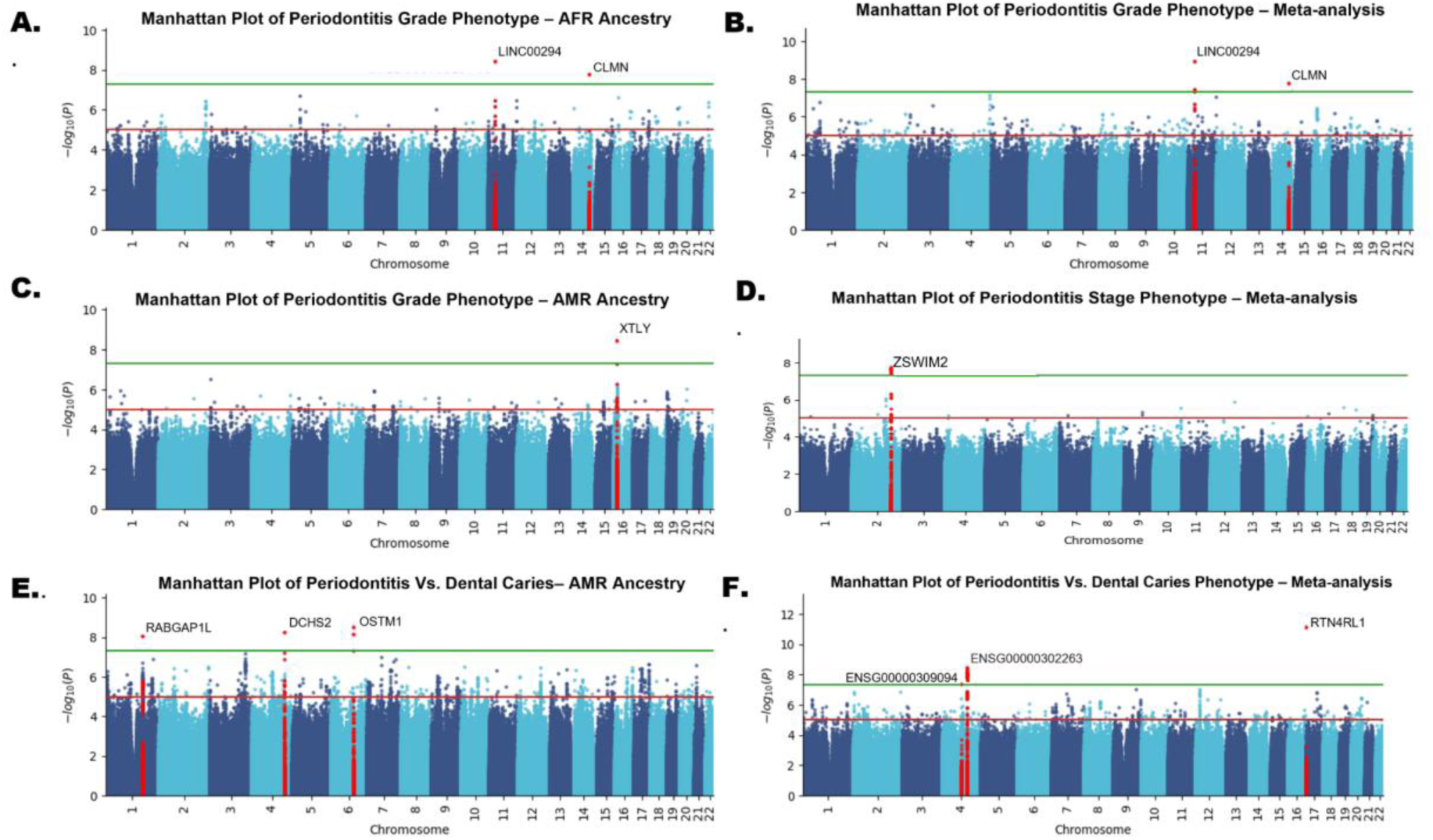
Manhattan plots for GWAS analysis with significant hits. Panels A (AFR grade), C (AMR grade), and E (AMR periodontitis vs. dental caries) show ancestry-specific results; panels B (grade), D (stage), and F (periodontitis vs. dental caries) show the corresponding meta-analyses. Each SNP’s -log^10^P is plotted by chromosome, with the green line denoting genome-wide significance (P = 5×10^-8^) and the red line the suggestive threshold (P = 1×10^-5^). The most significant loci in each analysis are labeled.

**Table 2.** Genome-wide significant ancestry-specific loci identified in phenotype-stratified GWAS.

| Ancestry | SNP | CHR | BP | Nearest Gene | EA | MAF | $\beta$ | P Value |
| --- | --- | --- | --- | --- | --- | --- | --- | --- |
| Periodontitis Grade Phenotype |  |  |  |  |  |  |  |  |
| AFR | rs73482834 | 11 | 33079319 | LINC00294 | G | 0.056 | -1.44 | 3.988e-09 |
| AFR | rs139217059 | 14 | 95223573 | CLMN | C | 0.015 | -2.44 | 1.764e-08 |
| AMR | rs147819514 | 16 | 17100380 | XYLT1 | AAT | 0.413 | 0.945 | 3.800e-09 |
| Periodontitis vs. Dental Caries |  |  |  |  |  |  |  |  |
| AMR | rs1183875194 | 1 | 174370463 | RABGAP1L | CTTT | 0.038 | 0.756 | 9.303e-09 |
| AMR | rs10677099 | 4 | 154251407 | DCHS2 | ATGTC | 0.097 | -0.505 | 5.889e-09 |
| AMR | rs113597243 | 6 | 108073100 | OSTM1 | A | 0.018 | 1.137 | 3.244e-09 |

SNP ID indicates the unique variant identifier; CHR indicates the chromosome number; BP indicates the base-pair position; EA indicates the effect allele tested; MAF indicates the minor allele frequency; $\beta$ indicates the per-allele effect size; P Value indicates the statistical significance of the association

Through the Meta-analysis of the ancestry-specific GWAS, we observed seven genome-wide significant loci across several of our study conditions (Figure 1). First, the meta-analysis identified three significant genome-wide loci for all periodontitis cases versus dental caries controls based on GWAS. These loci are ***RTN4RL1*** (rs1555520650, p value = 7.953e-12)**, *ENSG00000302263*** (rs62323100, p value = 3.738e-09), and ***ENSG00000302263*** (rs62323100, p value = 3.738e-09). Second, for the periodontitis stage phenotype, the meta-analysis identified ***ENSG00000295274*** (rs71303827, p-value = 1.657e-08), and for the periodontitis extent phenotype, the meta-analysis identified ***ZSWIM2*** (rs148253328, p-value = 1.411e-08). Last but not least, the meta-analysis identified two loci for the periodontitis grade phenotype, ***LINC00294*** (rs73482834, p-value = 1.227e-09) and ***CLMN*** (rs139217059, p-value = 1.764e-08), which also appeared in the AFR ancestry grade phenotype GWAS. We summarized the Meta-analysis findings, including summary statistics and variant annotations for each SNP in Table 3. Manhattan plots showing significant hits appear in Figure 1, and all GWAS and meta-analysis Manhattan plots, along with their QQ plots, are available in the supplementary material.

**Table 3.**
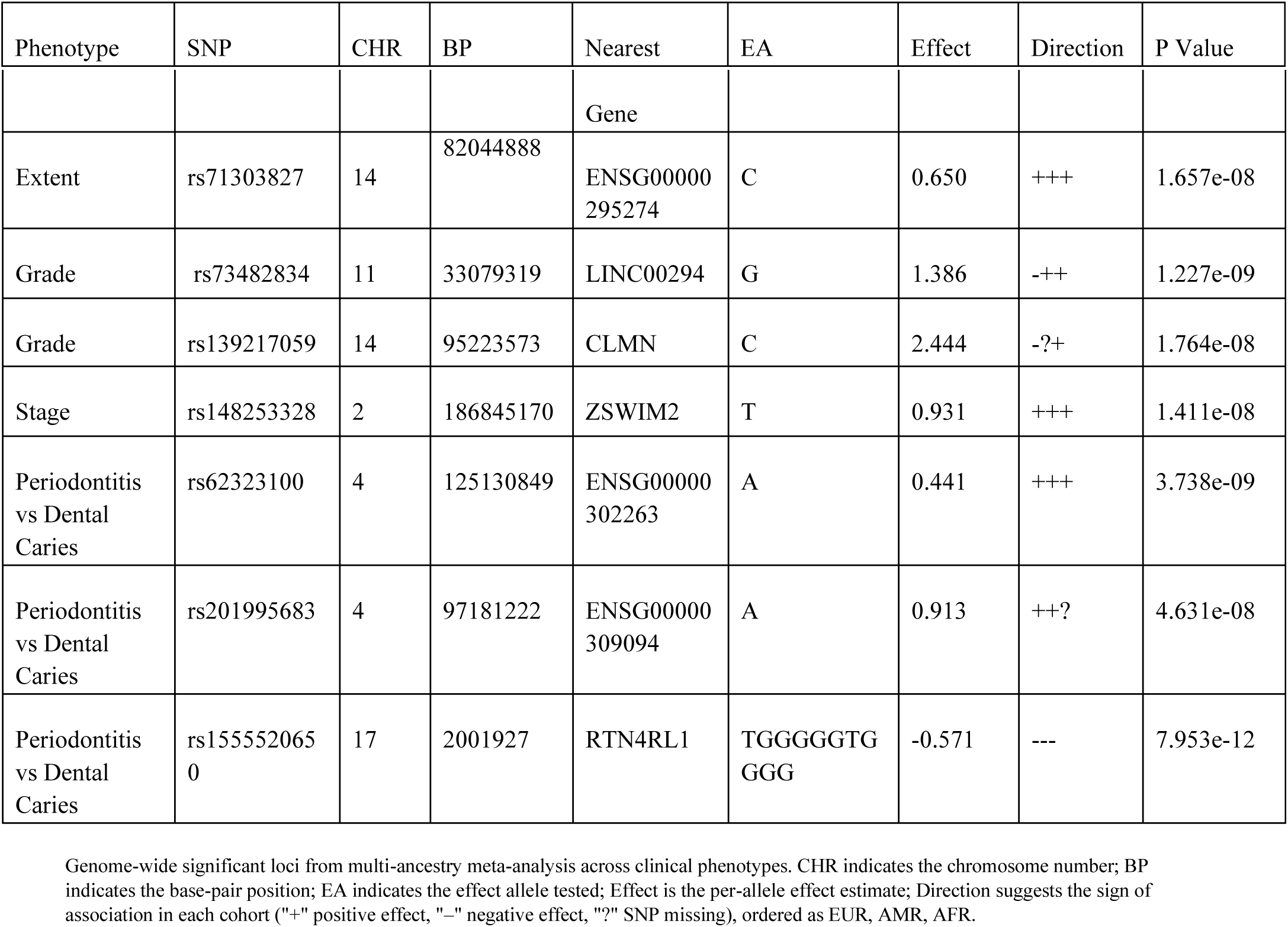
Genome-wide significant loci from multi-ancestry meta-analysis across clinical phenotypes.

### Regional Plot Analysis

In our post-GWAS analysis, we developed regional association plots to examine local linkage disequilibrium (LD) patterns and refine resolution of the signals at genome-wide significant loci, with the lead SNPs surpassing the significance threshold (p < 5×10⁻⁸). Our significant findings for periodontitis grade phenotype GWAS, we identified a genome-wide significant locus on chromosome 11 (lead SNP on *LINC00294*, ∼ 6 kb of *CSTF3* and *TCP11L1*, rs73482834, p = 3.988e-09, Fig. 2A) and chromosome 14 (lead SNP on *CLMN*, rs139217059, p = 1.764e-08, Fig. 2C) in the AMR study cohort. The lead SNP on chromosome 11 exhibits moderate (r² > 0.6) LD that extends across 33.0–33.2 Mb, indicating a focused association peak in this region. The lead SNP on chromosome 14 associates a moderate LD concentrated between 95.2–95.3 Mb, suggesting a tightly clustered signal. In the AFR study population, chromosome 16 (lead SNP on *XYLT1*, rs147819514, p = 3.800e-09, Fig. 2E) has multiple surrounding LD signals centered around 17.1–17.2 Mb. In our periodontitis vs. dental caries GWAS, we observed three genome-wide significant signals in our AMR cohort. Chromosome 1 genome-wide significant locus was observed (lead SNP on near *RABGAP1L*, rs1183875194, p = 9.303e-09, Fig. 2B), with moderate LD distributed around 174.2–174.6 Mb. On chromosome 4, we observed a signal (Lead SNP on *DCHS2*, rs10677099, p = 5.889e-09, Fig. 2F), with strong LD (r² > 0.8) concentrated between 154.25–154.30 Mb. Chromosome 6 (*OSTM1*, rs113597243, p = 3.244e-09, Fig. 2F), with the moderate and strong LD centered around 108.0–108.1 Mb.

**Figure 2.**
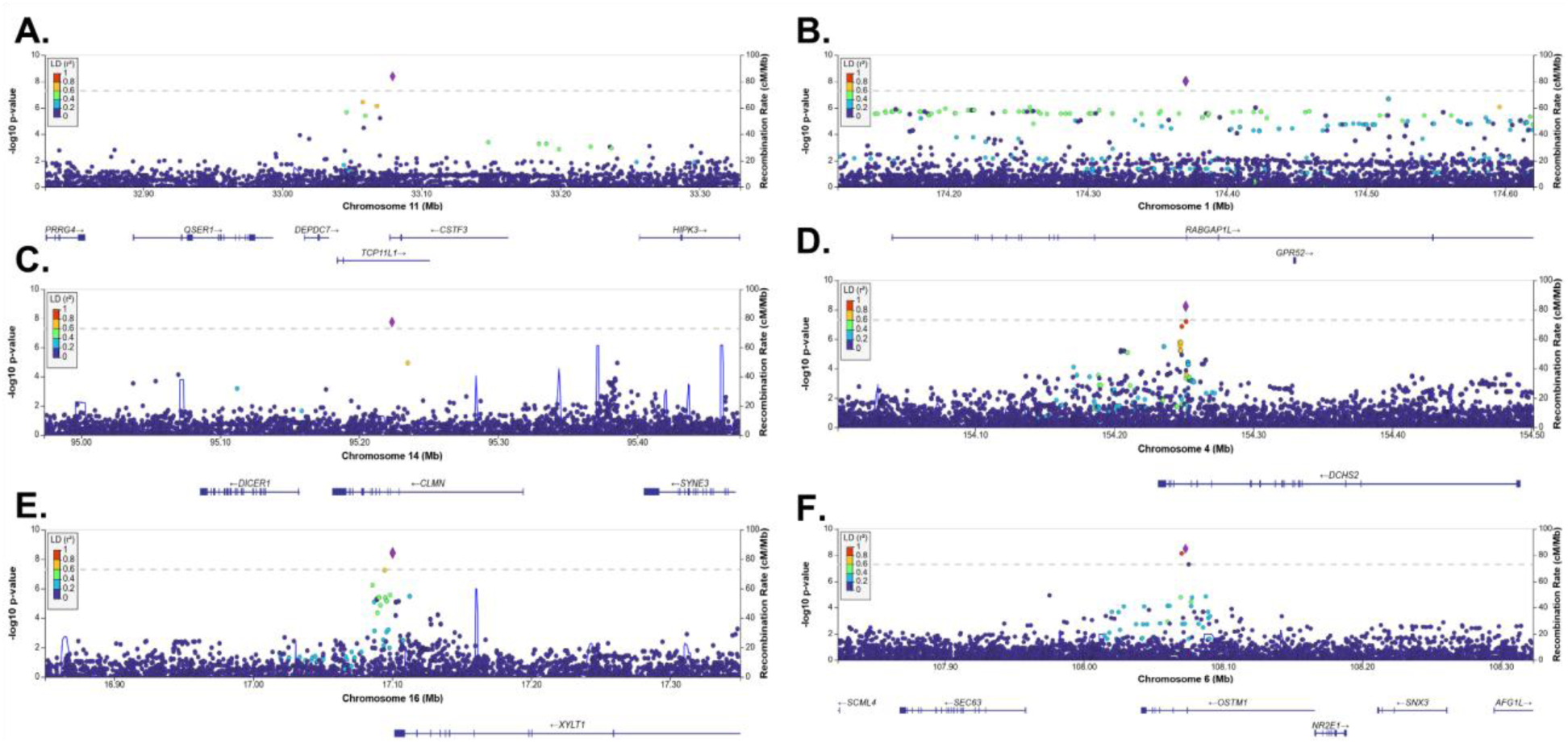
Regional association plots produced with LocusZoom illustrate the six loci with the strongest (P < 5 × 10^−8^) association signals in the periodontitis-based GWAS series. The position on the X-axis corresponds to genomic position, and the position on the Y-axis corresponds to each SNP’s –log_10_(*P* values). Each circle illustrates an SNP tested for association with our periodontitis phenotype. The lead SNP is highlighted in purple, while other variants are color-coded by their r^2^, a measure of LD, against the lead SNP. Figure 2A and 2C correspond with our periodontitis grade-based GWAS in AFR populations (n = 360). Figure 2E corresponds with our periodontitis grade-based GWAS in the AMR population (n= 404). Figures 2B, 2D, and 2F are associated with our periodontitis vs. dental caries phenotype in the AMR population (n = 3409).

### Periodontitis vs. Dental Caries Associated Genes

*OSTM1* was identified as a significant locus in our periodontitis vs dental caries GWAS analysis for AMR ancestry. *OSTM1* encodes a type I transmembrane protein indispensable for osteoclast differentiation and functional bone resorption [19]. Osteoclasts are specialized, multinucleated monocyte/macrophage lineage cells that adhere to bone surfaces and secrete proteolytic enzymes to dissolve mineralized matrices, thereby driving physiological and pathological bone remodeling [20]. *OSTM1* is highly expressed in mature osteoclasts and across multiple hematopoietic lineages, including activated macrophages, where it mediates the intercellular crosstalk required for fully functional osteoclastogenesis [20]. Another gene identified in our periodontitis vs dental caries GWAS for AMR ancestry is *DCHS2*, which produces a cadherin-related protein involved in bone mineralization and tissue morphogenesis. Although *DCHS2* has not been directly linked to periodontitis or related oral diseases, prior studies have associated it with craniofacial morphology and cartilage differentiation [21], especially in Latin American populations genetically similar to our admixed American cohort, where this gene showed significant association. The identification of *DCHS2* as a substantial locus in our admixed American ancestry GWAS supports its potential relevance in population-specific craniofacial and periodontal diseases. The third gene identified in the AMR ancestry periodontitis vs dental caries GWAS is *RABGAP1L*, which is described as an encoder for a GTPase-activating protein that modulates Rab7A and Rab10, corresponding to vesicle trafficking and antimicrobial autophagy [22]. Its part in cell immunity may suggest its potential importance in bacterial clearance and inflammatory regulation, a process relevant to periodontitis disease progression. The multi-ancestry meta-analysis for the periodontitis vs. dental caries GWAS indicated the gene *RTN4RL1*, a gene associated with cell-matrix adhesion in cementoblasts. A prior study suggested *RTN4RL1* plays a key role in the cementum–ECM interface genes [23]. Functional enrichment performed in the study revealed that ECM binding (GO:0050840) and cell adhesion molecule binding (GO:0050839) were dominant molecular functions in cementoblastic cells, implicating that the dysregulation of *RTN4RL1* could compromise cementum integrity and exacerbate periodontal breakdown under inflammatory conditions. Although no studies have directly associated *RTN4RL1* with periodontal diseases, our results advocate its continued exploration in this context.

### Periodontitis Grade Associated Genes

*XYLT1* was a significant gene associated with grade phenotype GWAS findings in the AMR ancestry. *XYLT1* encodes xylosyltransferase-I, the enzyme that catalyzes the initial step in proteoglycan assembly, which is crucial for extracellular matrix formation and corresponding structure in connective tissues and skeletal structures [24]. Inflammatory cytokines have been shown to alter *XYLT1* expression in fibroblasts, linking it functionally to tissue remodeling during inflammation [25,26]. Therefore, *XYLT1* is a compelling candidate for future exploration in periodontitis and related oral pathologies. *LINC00294* is one of the two significant genes appearing in the AFR ancestry grade GWAS and reinforced by the meta-analysis, suggesting a multi-ancestry association with aggressive versus chronic periodontitis. While not directly related to periodontitis, *LINC00294* has been shown to modulate inflammatory and mitochondrial stress responses in endothelial and glioma cancer cell models [27]. Its upregulation reduces pro-inflammatory cytokine production, aligning with biological processes relevant to periodontal tissue integrity [27]. These findings suggest that *LINC00294* may play a broader immunomodulatory role, indicating a potential molecular association with systemic inflammation. The other loci with specific and multi-ancestry association were *CLMN,* which encodes a calponin-homology domain-containing protein involved in cytoskeletal regulation, cell cycle regulation, and neuronal differentiation, named Calmin [28]. Notably, *CLMN* has been identified as a vitamin D-responsive gene. Prior studies have explored the association between vitamin D deficiency and periodontitis [29], suggesting that *CLMN* may function as a downstream effector of vitamin D’s immunomodulatory roles in oral tissues. Furthermore, its retinoic acid-dependent expression and regulatory functions in cell differentiation indicate an expanded relevance to inflammatory diseases such as rheumatoid arthritis, where immune dysfunction and epithelial integrity are recognized pathological features [30].

## Discussion

Using the WGS data available for more than 3800 periodontitis patients in the AOURP researcher workbench, our GWAS analysis pipeline uncovered 10 loci associated with periodontitis and its sub-phenotypes. We identified six ancestry-specific loci, three of which differentiate the association with periodontitis from dental caries. A signal of note is *OSTM1*, which, in a prior study performed by Pata, He’raud, & Vacher, suggests that loss-of-function mutations or downregulation of *OSTM1* may lead to severe autosomal-recessive osteopetrosis, characterized by craniofacial and alveolar bone abnormalities [19]. Since alveolar bone homeostasis depends on balanced osteoclast-mediated resorption and osteoblast-driven formation, the disruption of *OSTM1* may compromise periodontal support [31]. The mechanisms guiding osteoclast differentiation have also been previously examined for association with periodontitis [32]. Moreover, a prior investigation of *OSTM1’*s broad expression in macrophages and lymphoid tissues suggests a potential link between immune activation and bone remodeling [33]. The other three loci appear in the periodontitis grade phenotype GWAS, differentiating the association between aggressive or acute periodontitis versus chronic periodontitis. Amongst them, we identified possible associations with *CLMN* and *LINC00249* genes, which were further reinforced by the multi-ancestry meta-analyses, suggesting the finding of novel genes in other study schemes. Our multi-ancestry approaches improved statistical power while preserving population-specific signals, reducing bias from over-representing European ancestry and enhancing relevance to global populations [34, 35]. Our lead SNP at 33,079,319 bp lies within *LINC00294* and sits ∼5.8 kilobases downstream of *TCP11L1* and ∼5.3 kilobases upstream of CSTF3 (Supplementary Fig. 5), motivating follow-up of these neighboring genes given the potential for shared regulatory elements. *CSTF3* encodes one of three cleavage stimulation factors (with *CSTF1* and *CSTF2*) that form the CSTF complex, which mediates polyadenylation and 3′ end cleavage of pre-mRNAs. While *CSTF3* has not been investigated in periodontitis, a prior study examined *CSTF2*, another component of the same complex, in blood and tissue samples but found no differential expression compared to controls. However, correlations with immune-related genes were noted [36]. Given the cooperative function of CSTF3 with CSTF2 and CSTF1 in the CSTF complex, additional exploration of CSTF3 may be warranted in the context of periodontal immune regulation. Our findings highlight the immune, inflammatory, and bone remodeling pathways underpinning periodontal disease, which can be further confirmed with future in vitro studies.

As described in our methods, patients with dental caries who were never diagnosed with periodontitis were considered our control group. Dental caries was selected to ensure controls had dental records and reduce misclassification due to missing data. It also provides an appropriate comparator, sharing diagnostic pathways and risk factors with periodontitis while remaining clinically distinct and lacking periodontal tissue destruction. Thus, this approach allowed us to capture genetic signals corresponding to clinically defined periodontitis characteristics rather than general oral disease susceptibility. Among these, *OSTM1*, a gene critical for osteoclast maturation and skeletal integrity, may influence periodontal support through its role in alveolar bone homeostasis [31,33]. *RABGAP1L*, implicated in tissue remodeling and immune regulation, merits further study in oral disease contexts [37]. This prior study recognized differentially expressed *RABGAP1L* in a transcriptomic comparison analysis of developing versus remodeling periodontium, suggesting a role in tissue remodeling and immune activation within the oral environment [36]. These findings nominate *RABGAP1L* for further study as a candidate gene involved in host-microbe interactions and periodontal tissue dynamics.

While our findings offer novel insights into the genetics of periodontitis, several methodological limitations warrant careful consideration. First, our phenotype definitions relied on retrospective EHR data collected from AOURP participants, which may be missing data context relevant to the narrowly focused dental care sector. However, without access to raw clinical records, our definitions of stage, grade, and extent relied on OMOP concepts and billing codes, which may not fully align with the 2017 WWC standards and could limit direct links between genetic associations and disease characteristics. Despite this ambiguity, genome-wide significant loci emerge, suggesting that our classification approximation approach still captures meaningful biological variation. Secondly, we have relatively small cohort sizes in some periodontitis phenotypes, particularly for the stage phenotype, especially when splitting by ancestry. Future work is warranted to use machine learning models trained on large data sources to infer the periodontitis 2017 WWC [7] classification, improve our current classification approximation method, and potentially increase cohort sizes. Lastly, another limitation can be observed in the interpretation of our results. Our findings suggest strong indirect literature evidence for inflammation and oral bone integrity rather than established periodontitis-specific roles. Future studies investigating a more direct link between periodontitis and the corresponding mechanisms may bolster our findings’ association with the disease. Moreover, our findings included locus *ZSWIM2*, *ENSG00000295274*, *ENSG00000309094*, and *ENSG00000302263*, which lacked direct characterization in periodontitis research and minimal evidence linking them to any disease phenotype, revealing a significant gap in broader genetic and clinical understanding. Moreover, the correlative findings of our studies have not been tested *in vitro* or *in vivo*, leaving the mechanistic roles of identified genes in periodontitis unconfirmed, which we suggest for future work.

The contributions of our study integrate EHR phenotyping and genetic discovery to advance understanding of periodontitis. Using OMOP-standardized codes, we stratified periodontitis by its complex clinically relevant dimensions, providing a novel framework for GWAS to uncover genetic associations linked to periodontitis severity, progression, and distribution that would be missed in aggregate analyses. The AOURP dataset has not been previously applied to investigate periodontitis or other oral diseases, positioning our study as a novel contribution to genomic oral health research. We also leveraged the periodontitis stage, grade, and extent phenotypes to identify periodontitis-associated candidate genes using the AOURP cohort. This approach identified loci consistent with prior studies, including our finding in *OSTM1* and *DCHS2*, whose roles in osteoclast regulation in bone remodeling and craniofacial morphology, respectively. Notably, our stratified, ancestry-specific, and multi-ancestry analyses revealed novel candidates such as *LINC00294* and *CLMN* that had not previously been implicated in periodontitis. These findings highlight the potential discovery of novel and concordant loci across heterogeneous ancestries, while broadening the genetic contributors associated with disease heterogeneity.

## Conclusion

Potential loci underlying periodontitis and its sub-phenotype heterogeneity were identified by integrating multi-ancestry GWAS with clinically stratified stage, grade, and extent phenotypes from the AOURP. The current approach overcomes the constraints of homogeneous cohorts and broad phenotype definitions, emphasizing how diverse populations can yield underexplored insights. While this study’s findings demonstrate a practical genomic foundation, functional and longitudinal studies are required to validate causal mechanisms and to evaluate their clinical relevance. This study establishes the groundwork for subsequent precision phenotyping in periodontal research by leveraging the rich, linked EHR and genomic data to uncover novel genetic insights, and the prospective incorporation of advanced information-extraction strategies could further amplify the clinical relevance of these findings.

## Materials and Methods

### Data Source

AOURP (v.8) is a U.S. longitudinal biomedical data repository with extensive multi-modal data from over 600,000 participants, with a significant focus on including diverse populations [16]. Utilizing a secure, cloud-based Researcher Workbench, researchers can access EHRs, detailed survey responses, physical measurements, biospecimens, and wearable sensor streams. Participants can consent to short-read whole-genome sequencing (srWGS), contributing to one of the largest multi-ancestry genomic datasets publicly accessible via the secure, cloud-based Researcher Workbench. AOURP v.8 includes the srWGS from over 400,000 participants. This rich and diverse resource forms the backbone of our GWAS analyses across multiple clinically stratified periodontitis phenotypes.

### EHR phenotyping and cohort definition

Since AOURP utilizes the OMOP common data model, patient diagnoses (conditions) are primarily available as standardized OMOP concepts. In most cases, the billing diagnosis codes shared by the data sharing partner (source) are also available. The standardized OMOP concept identifiers map to predefined standard Systematized Nomenclature of Medicine - Clinical Terms (SNOMED-CT) codes [38], and the source billing codes are in the International Classification of Diseases ninth and tenth revisions (ICD-9, ICD-10) standards. Therefore, our analysis identified our cohorts using standard and source concepts, as shown in Supplementary Table 1. Although the standard for dental care is to document the dental diagnosis in SNODENT [39] and the current best practice for periodontitis classification is following the 2017 WWC, those specific codes are not currently available in the AOURP. However, the available standardized and billing codes still include whether the periodontitis is aggressive or chronic, along with severity and extent classification [39]. Therefore, we decided to classify the diagnosis following the stage, grade, and extent principles: stage (slight, moderate, severe), grade (chronic, acute, or aggressive), and extent (localized, generalized).

Given that not all participants’ dental records may have been shared along with their EHRs, we cannot assume that patients who did not have a periodontitis diagnosis are non-periodontitis patients. Therefore, we decided only to consider patients with dental diagnoses, specifically dental caries, in our control group. In our periodontitis vs. dental caries GWAS, our cases were patients with any periodontitis diagnosis, and our controls were patients with dental caries who were never diagnosed with any type of periodontitis. Furthermore, in periodontitis phenotype subgroup GWAS, only patients with specified classifications were included. For the extent phenotype, cases were defined as generalized periodontitis and controls as localized periodontitis (Table 1). Patients lacking a defined extent classification were excluded. The analysis did not include patients without a label in the extent category. These decisions aimed to minimize the risk of misclassification by ensuring participants were assigned only to diagnoses that reflected their clinical records.

All patients included in our analysis should have shared their srWGS. AOURP computed relatedness using pairwise kinship coefficients. Coefficients among pairs with a score of 0.1 or higher were suggested as first-degree relatives and were excluded from the GWAS analysis. Additional exclusion criteria included participants whose earliest diagnosis of periodontitis or dental caries was under the age of 18.

### Genome-Wide Association Study

We conducted four genome-wide association studies (GWAS) in this study. The first was for periodontitis condition in general, regardless of its classification (subphenotype), and three subphenotype-specific periodontitis GWAS, stratified by disease stage, grade, and extent. The phenotypic and sub-phenotypic features were extracted from the AOURP EHR data as described in the study cohort section above, and we binarized them for the GWAS analysis. For the general periodontitis GWAS, cases included patients diagnosed with periodontitis, and controls included patients diagnosed with dental caries. To ensure distinct groups, individuals with both conditions were excluded from both cohorts. For the stage, we defined our cases as patients with severe periodontitis and our controls as patients with slight or moderate periodontitis. For the grade sub-phenotype, we compared individuals classified as acute or aggressive versus chronic periodontitis and, for extent, generalized versus localized periodontitis. Our analysis design enabled us to discover genetic loci associated with intra-disease heterogeneity and simultaneously assess the specificity of these associations against related oral pathologies.

The GWAS analysis utilized PLINK v1.9 to perform logistic regression adjusted for sex, age, self-reported ethnicity, and 16 principal components. PLINK parameters include: Single Nucleotide Polymorphisms (SNPs) with small genotyping rates (--geno 0.05), SNPs with low minor allele frequency (--maf 0.01), and subjects with high amounts of genotype missingness (--mind 0.2). AOURP provided genetically inferred ancestry fractions for each participant utilizing Rye [41], a rapid PCA-based algorithm alongside reference panels from the 1000 Genomes Project and Human Genome Diversity Project. We extracted the assigned ancestry label from each sample and applied these groupings to perform separate ancestry-based GWAS analyses. For this study, we focused our multi-ancestry analyses on participants with predominant African (AFR), European (EUR), and Admixed American (AMR) ancestry groups, as these represented the most highly populated continental ancestries in our cohort. The genome-wide significance threshold was set at p < 5 × 10^-8^ reflecting a Bonferroni correction, a standard widely accepted for minimizing false-positive associations in GWAS.

The AFR, AMR, and EUR GWAS summary results were aggregated to conduct our meta-analysis utilizing METAL [42], an inverse-variance fixed-effects model. Inputs included β values, standard errors, and N per variant per ancestry group. We also performed Cochran’s Q-test to detect ancestry-specific heterogeneity. This method ensures that large ancestry groups contribute appropriately without suppressing population-specific signals.

## Supporting information

Supplement Figure 1. Periodontitis vs Dental caries GWAS Manhattan plots

Supplement Figure 2. Periodontitis Extent Phenotype GWAS Manhattan plots

Supplement Figure 3. Periodontitis Grade Phenotype GWAS Manhattan plots

Supplement Figure 4. Periodontitis Stage Phenotype GWAS Manhattan plots

Supplement Table 1.Summary of clinical phenotype groupings

Supplement Figure 5. UCSC Genome Browser highlighting LINC00294

## Author Contributions

LR, CL, and DZ initialized the conceptualization of the project. KS, AN, DZ, and LR designed the methods. KS, CL, and LR defined the phenotypes and identified the relevant cohort. KS conducted the formal analysis with substantial input from AN, DZ, and YH. KS, AN, LR, and DZ interpreted the results. KS, CL, and LR led the writing of the first draft with substantial inputs from JI and YH. Cl, JI, BT, and MW reviewed the dental aspects. LR and DZ supervised the execution of the project.

## Conflict of Interest Statement

The authors disclosed that no potential conflicts of interest

## Ethics Statement

The All of Us protocol was reviewed and approved by the Institutional Review Board (IRB) of the All of Us Research Program (2021-02-TN-001). The All of Us IRB follows the regulations and guidance of the NIH Office for Human Research Protections for all studies, ensuring that the rights and welfare of research participants are overseen and protected uniformly. The present study did not directly involve participants, only their data; and the data available in the Researcher Workbench has been carefully checked and altered to remove identifying information while preserving its scientific utility. The Researcher Workbench employs a data passport model, through which authorized users do not need IRB review for each research project. As such, this research is considered non-human subjects research, and project-specific IRB approvals or waivers are not required. In compliance with the All of Us Data and Statistics Dissemination Policy, no results that include fewer than 20 participants are reported, in order to protect participant privacy.

## Funding Statement

Research reported in this publication was supported by the National Library of Medicine of the National Institutes of Health under Award Number R01LM014249, and by the National Institute of Allergy and Infectious Diseases of the National Institutes of Health under Award Number R01AI175699. The content is solely the responsibility of the authors and does not necessarily represent the official views of the National Institutes of Health.

## Acknowledgments

We thank the participants of the All of Us Research Program for their invaluable contributions, which made this research possible. We also acknowledge the NIH *All of Us Research Program* for supporting research through its commitment to data accessibility.

## Data availability

The GWAS summary statistics developed in this study are derived from data from AOURP and will be available in coordination with AOURP. The patient-level demographic and genotype data can be accessed upon application request from the All of Us Research Program (https://www.researchallofus.org).

## Supplementary Files

**Supplementary Figure 1.**
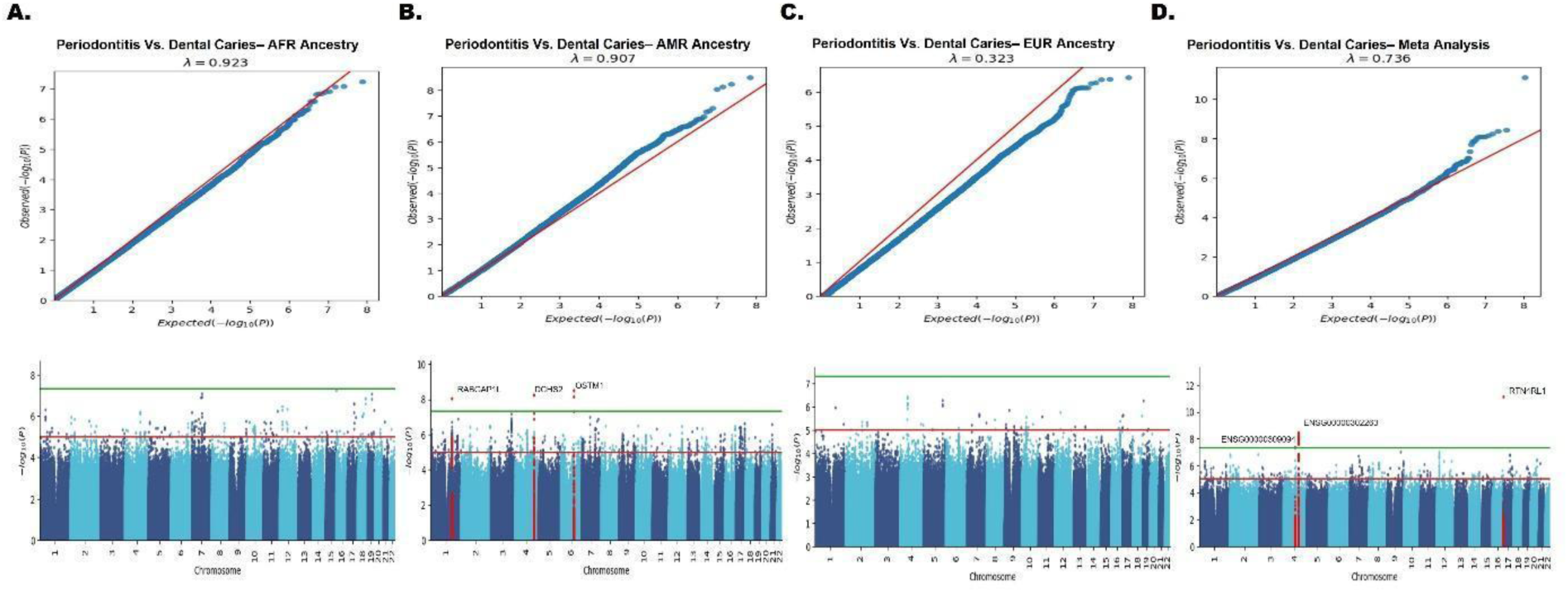
QQ plots and Manhattan plots for Periodontitis vs. Dental Caries GWAS. Each panel displays ancestry-stratified (A–C) and meta-analysis (D) results. The top row shows QQ plots comparing observed and expected –log₁₀(p) values with genomic inflation factor (λ). The bottom row shows Manhattan plots of –log₁₀(p) values by chromosomal position, with marked genome-wide and suggestive significance thresholds.

**Supplementary Figure 2.**
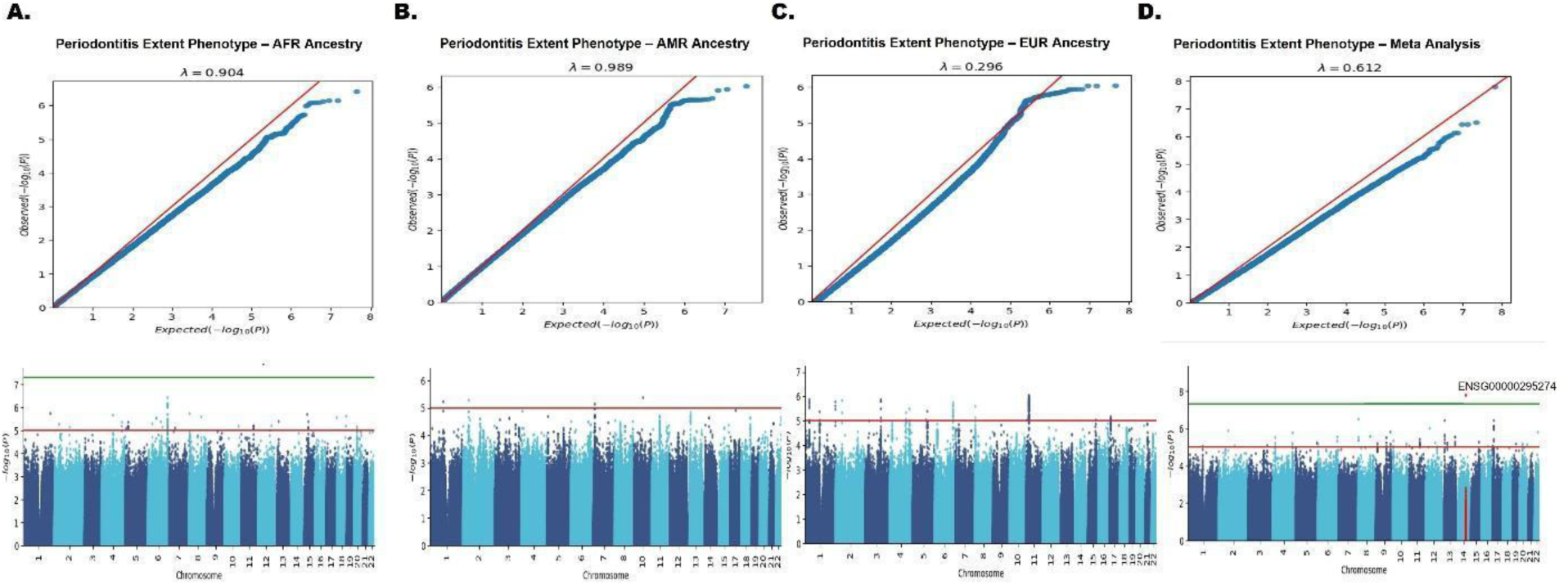
QQ plots and Manhattan plots for the Periodontitis Extent Phenotype GWAS. Panels A–C represent ancestry-stratified results; panel D shows the meta-analysis. QQ plots (top row) display genomic inflation, while Manhattan plots (bottom row) show association signals across chromosomes. Standard significance thresholds are indicated.

**Supplementary Figure 3.**
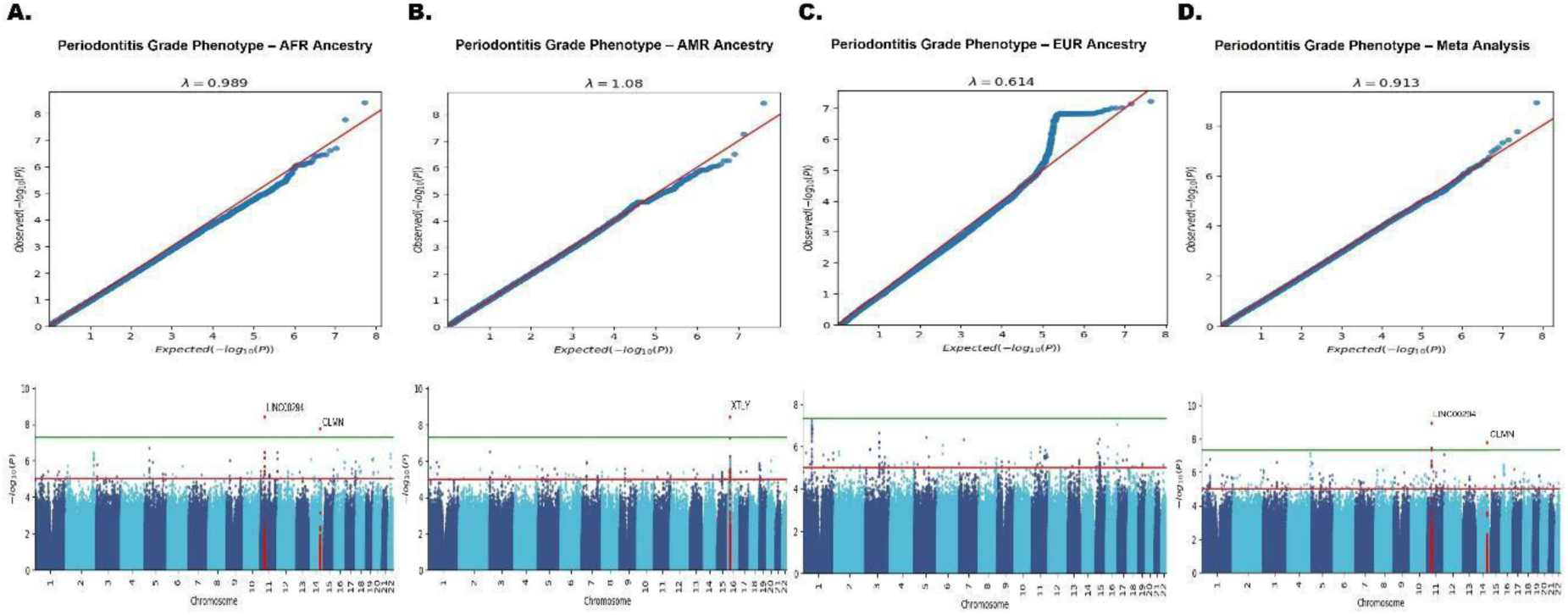
QQ plots and Manhattan plots for the Periodontitis Grade Phenotype GWAS. Panels A–C represent ancestry-stratified results; panel D shows the meta-analysis. QQ plots (top row) display genomic inflation, while Manhattan plots (bottom row) show association signals across chromosomes. Standard significance thresholds are indicated.

**Supplementary Figure 4.**
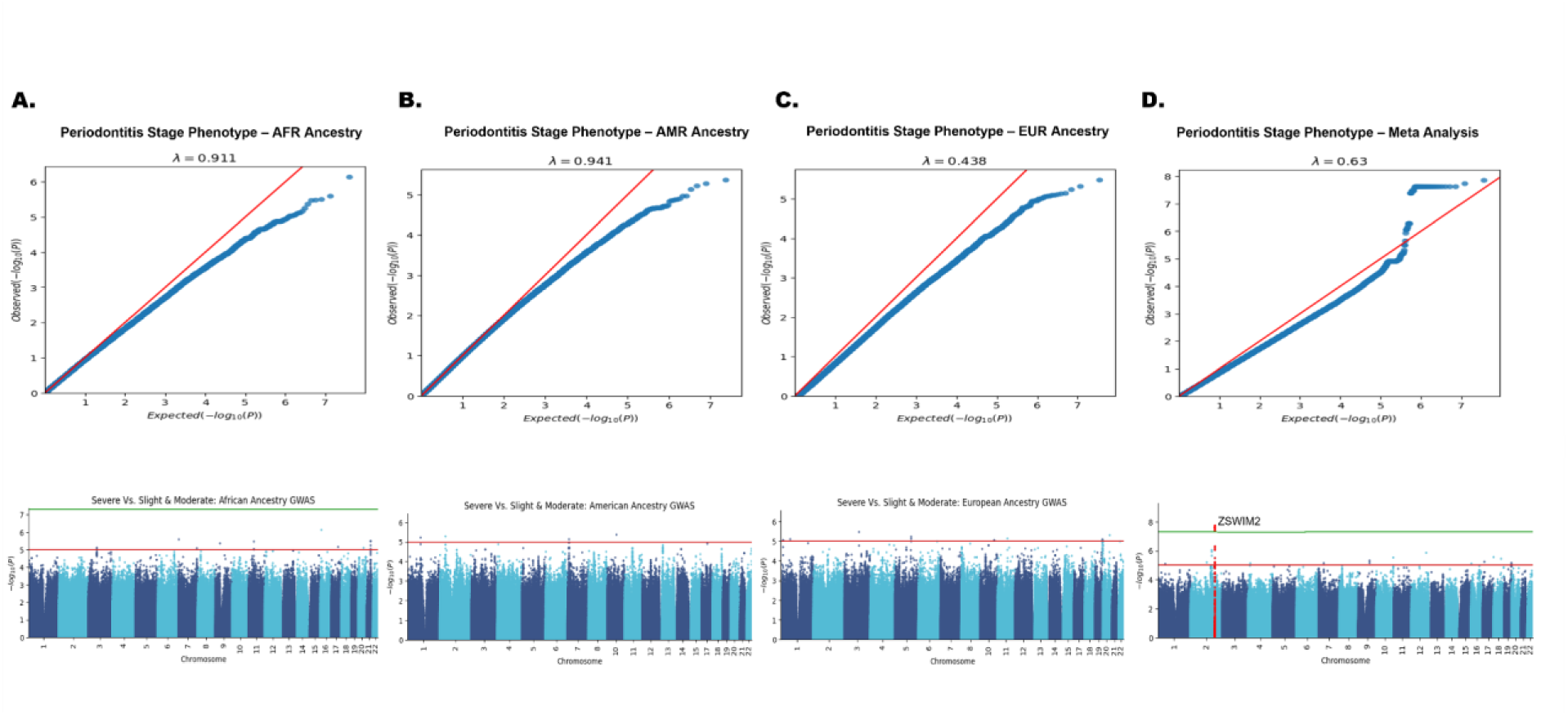
QQ plots and Manhattan plots for the Periodontitis Stage Phenotype GWAS. Panels A–C represent ancestry-stratified results; panel D shows the meta-analysis. QQ plots (top row) display genomic inflation, while Manhattan plots (bottom row) show association signals across chromosomes. Standard significance thresholds are indicated.

**Supplementary Figure 5.**
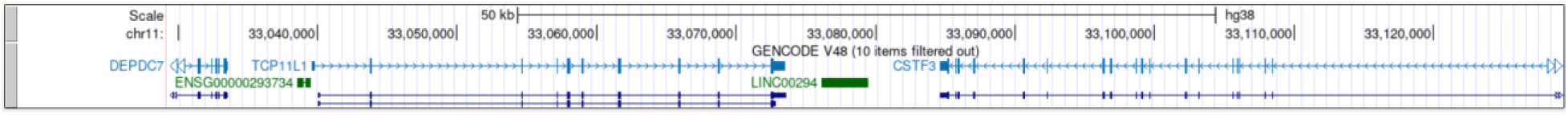
UCSC genome browser(Hg38) highlighting GWAS-identified site in LINC00294 with proximity to TCP11L1 and CSTF3. Our GWAS of periodontitis grade criteria identified a significant site within the LINC00294 locus. This site lies ∼5.7 kb downstream of TCP11L1 and ∼5.3 kb upstream of CSTF3.

**Supplementary Table 1.**
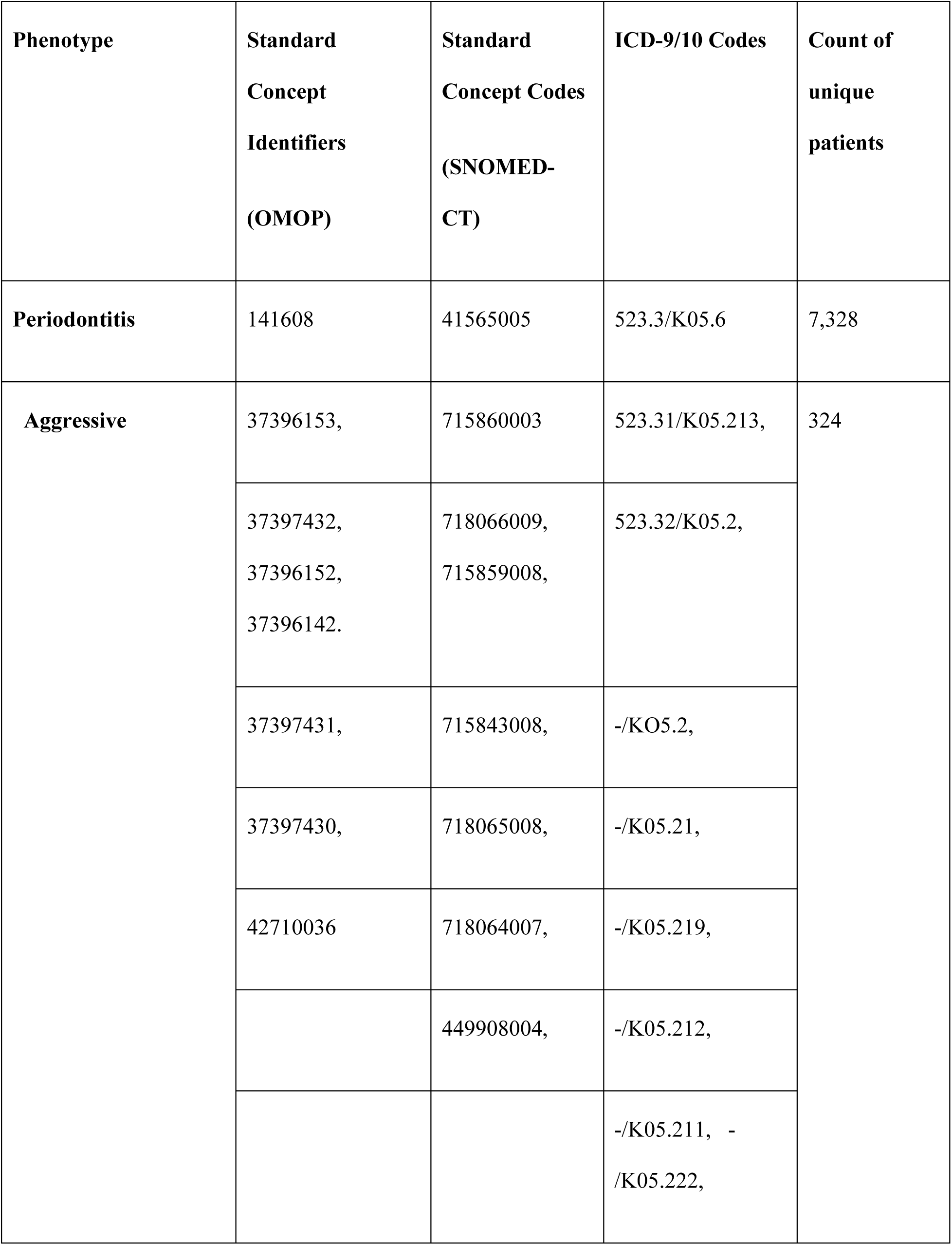
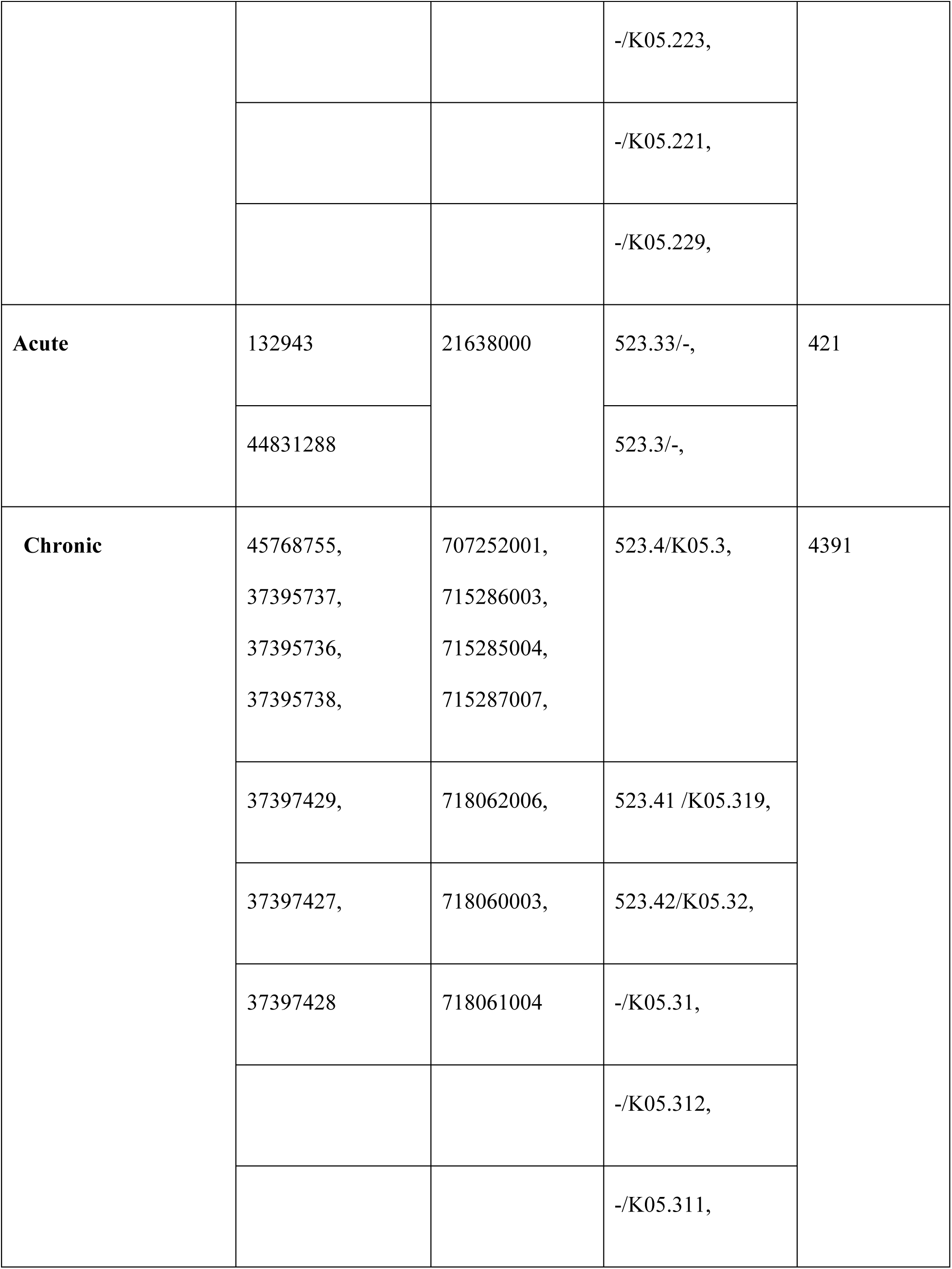
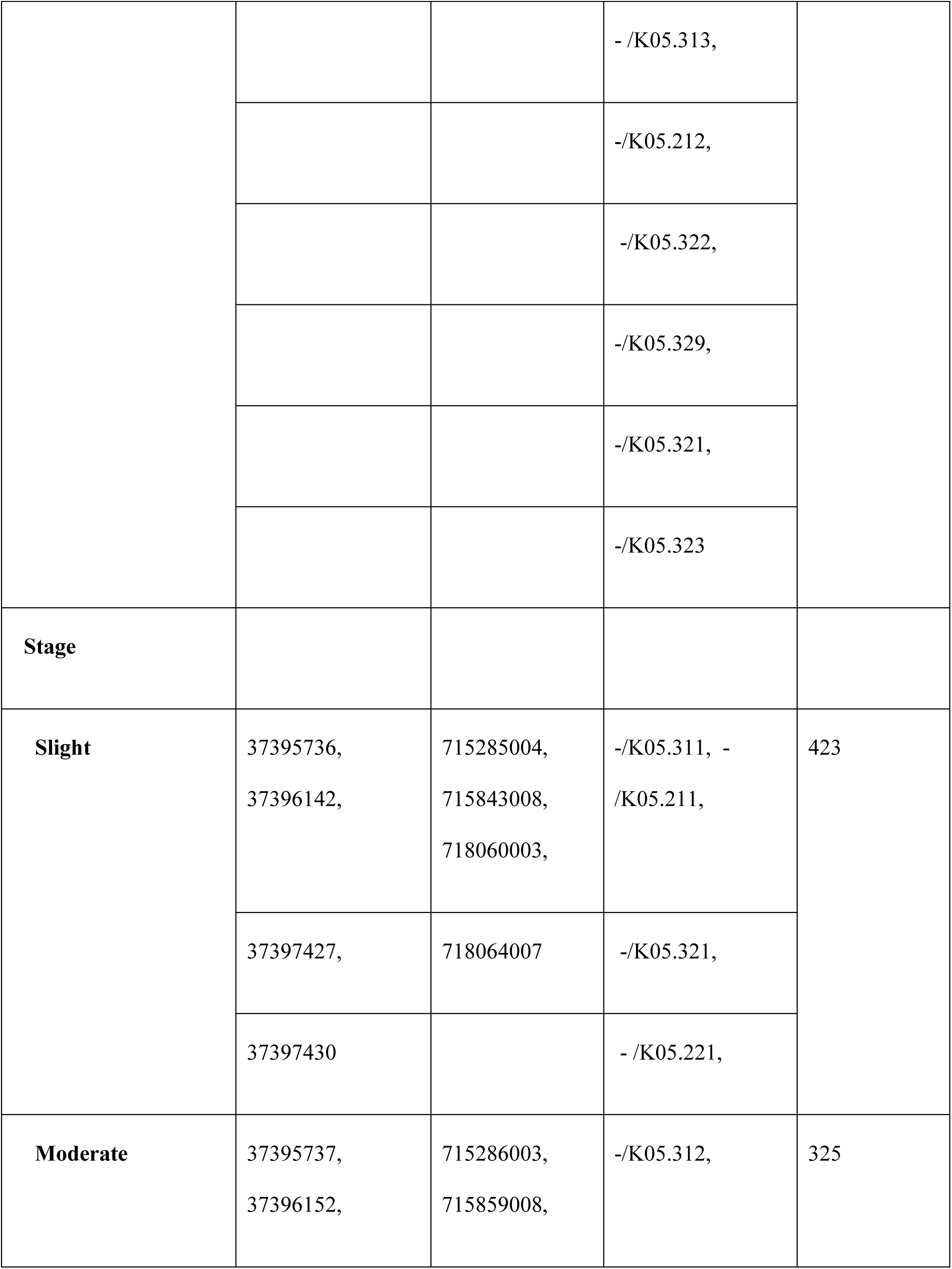
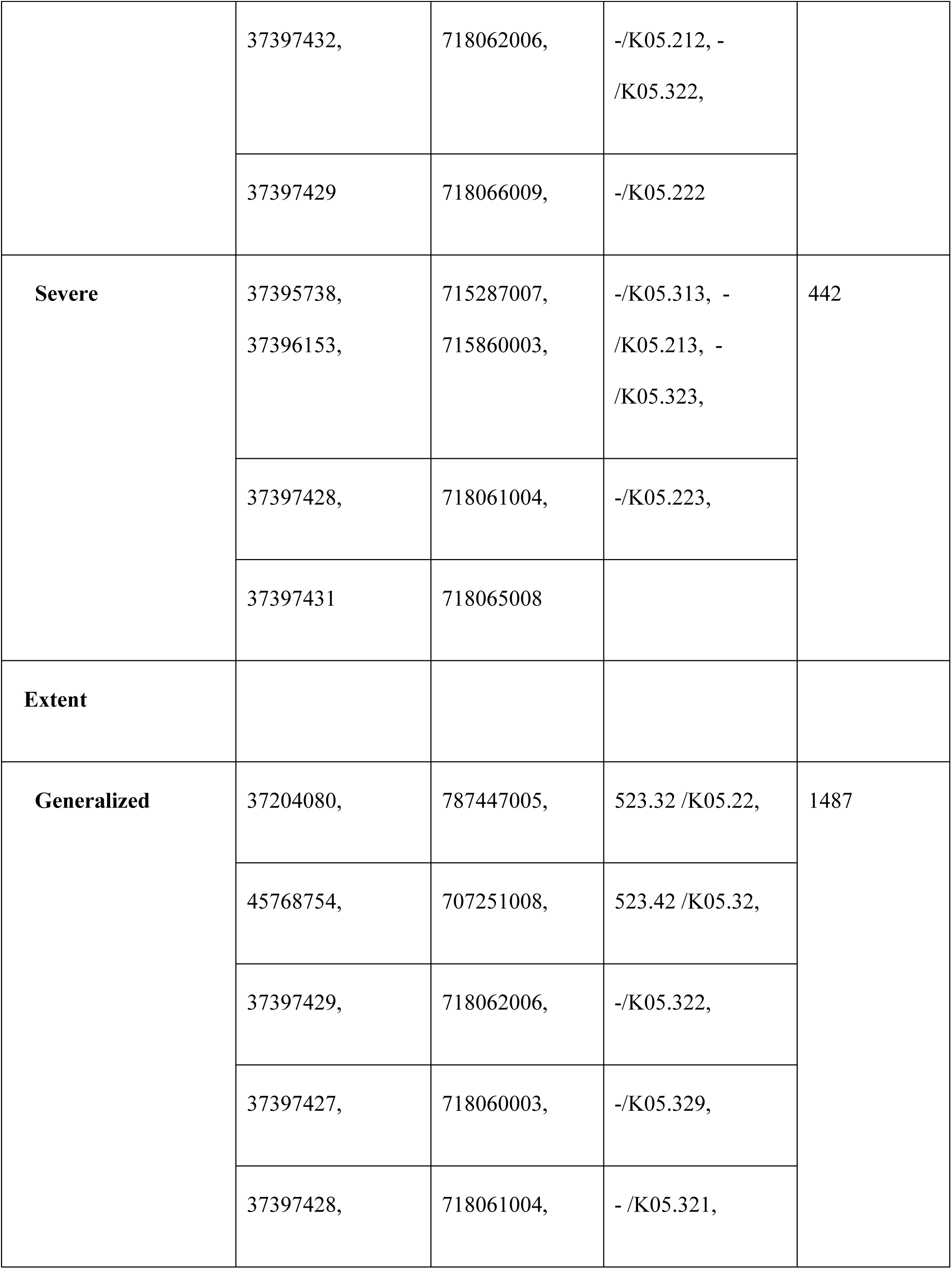
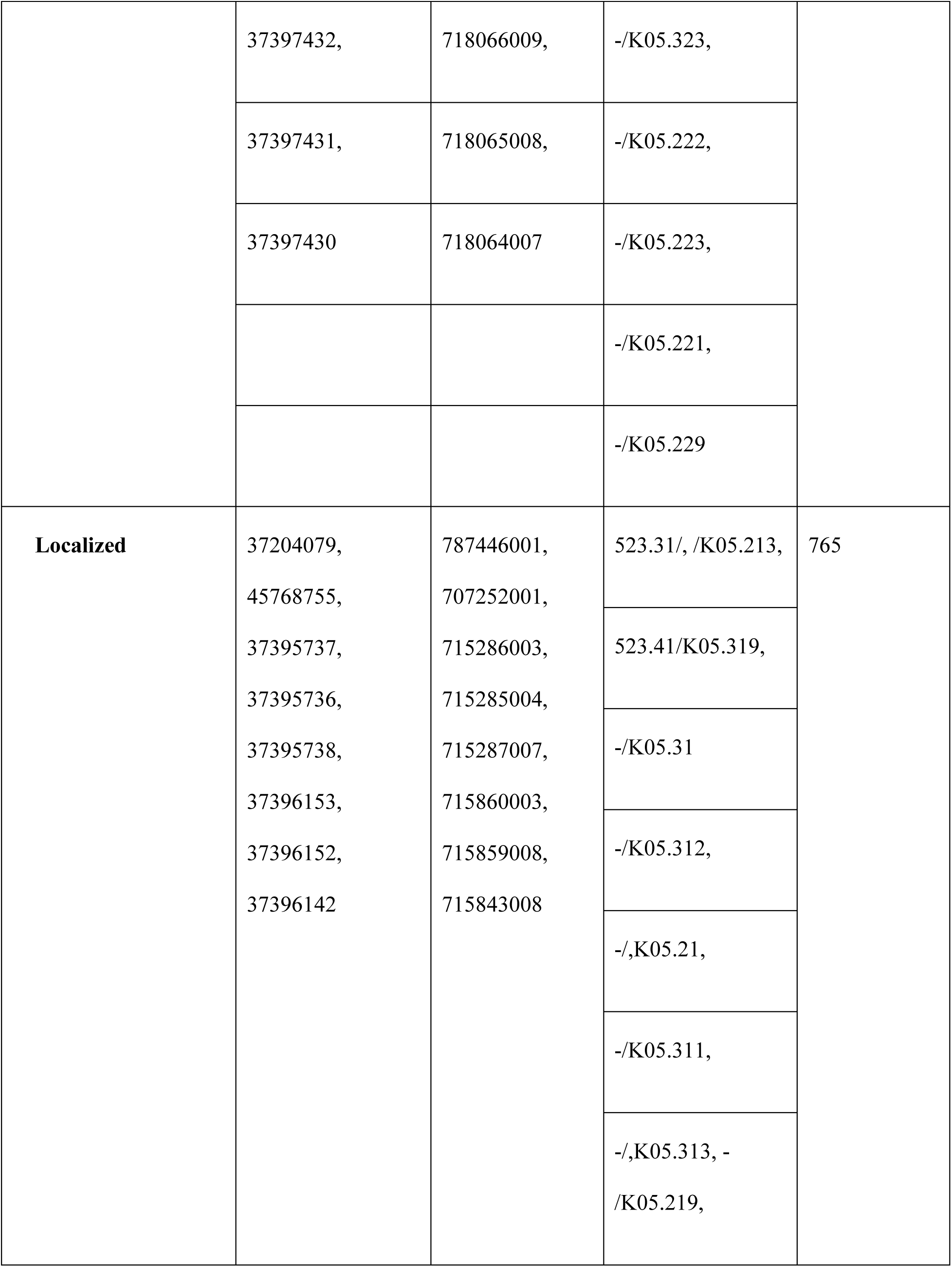
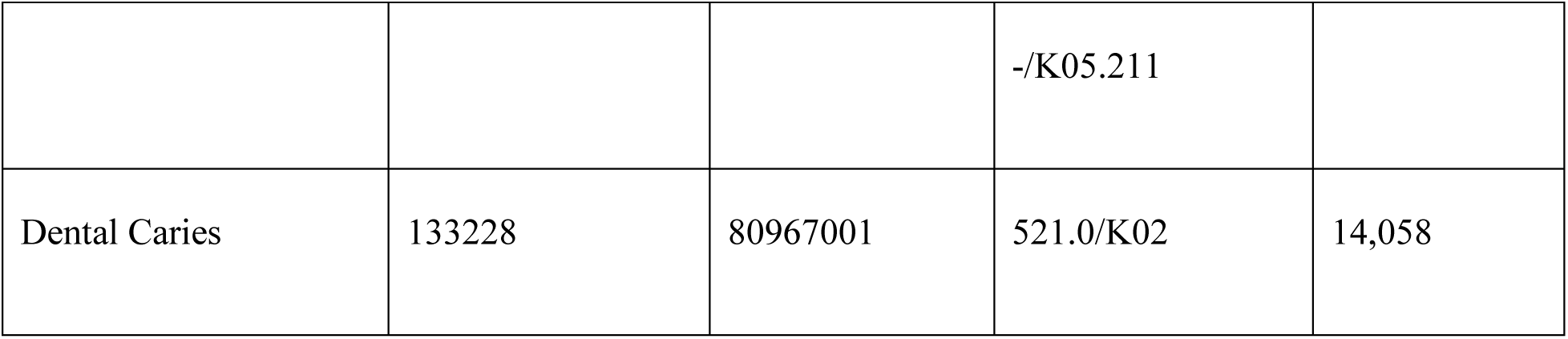
Summary of clinical phenotype groupings, associated OMOP concept identifiers, SNOMED-CT and ICD-9/10 codes, and the count of unique patients per phenotype. This table outlines the mapping structure used for defining periodontitis-related conditions in the analysis.

## Notes

### Competing Interest Statement

The authors have declared no competing interest.

