## Supplementary figures and images for "GWAS for Periodontitis Phenotypes Using Multi-Ancestry All of Us Research Platform"

### Supplement Figure 1. Periodontitis vs Dental caries GWAS Manhattan plots

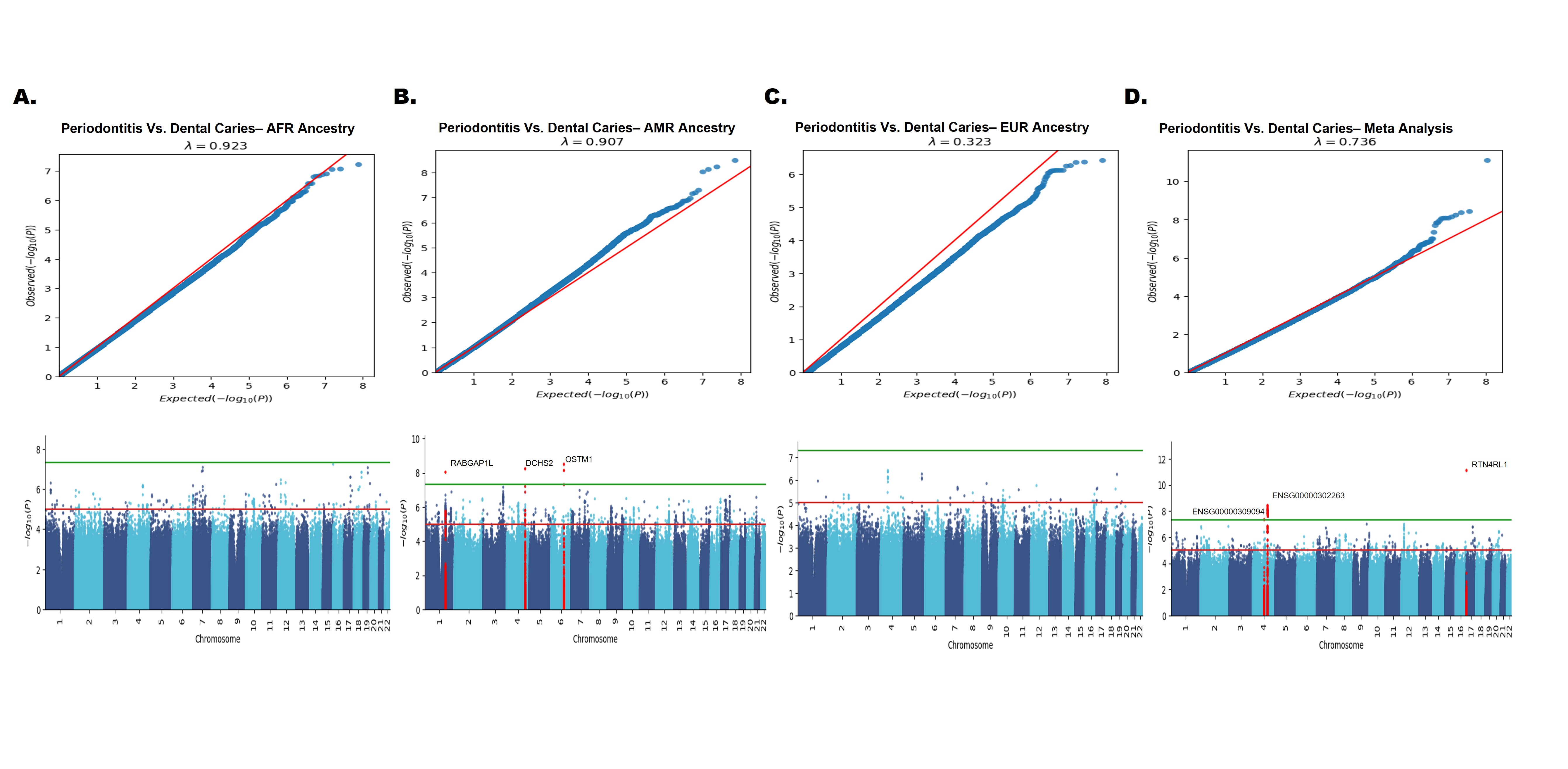

### Supplement Figure 2. Periodontitis Extent Phenotype GWAS Manhattan plots

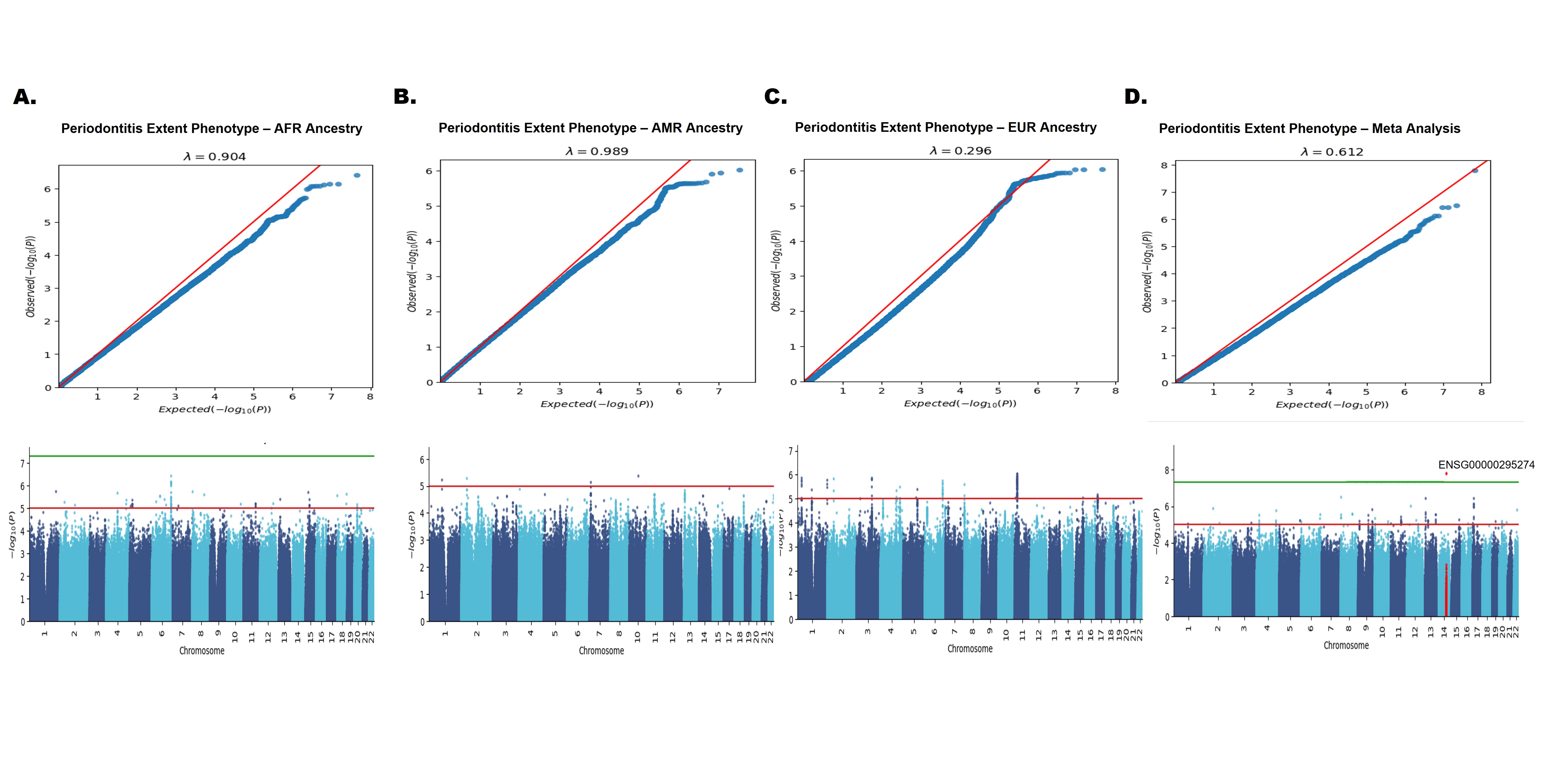

### Supplement Figure 3. Periodontitis Grade Phenotype GWAS Manhattan plots

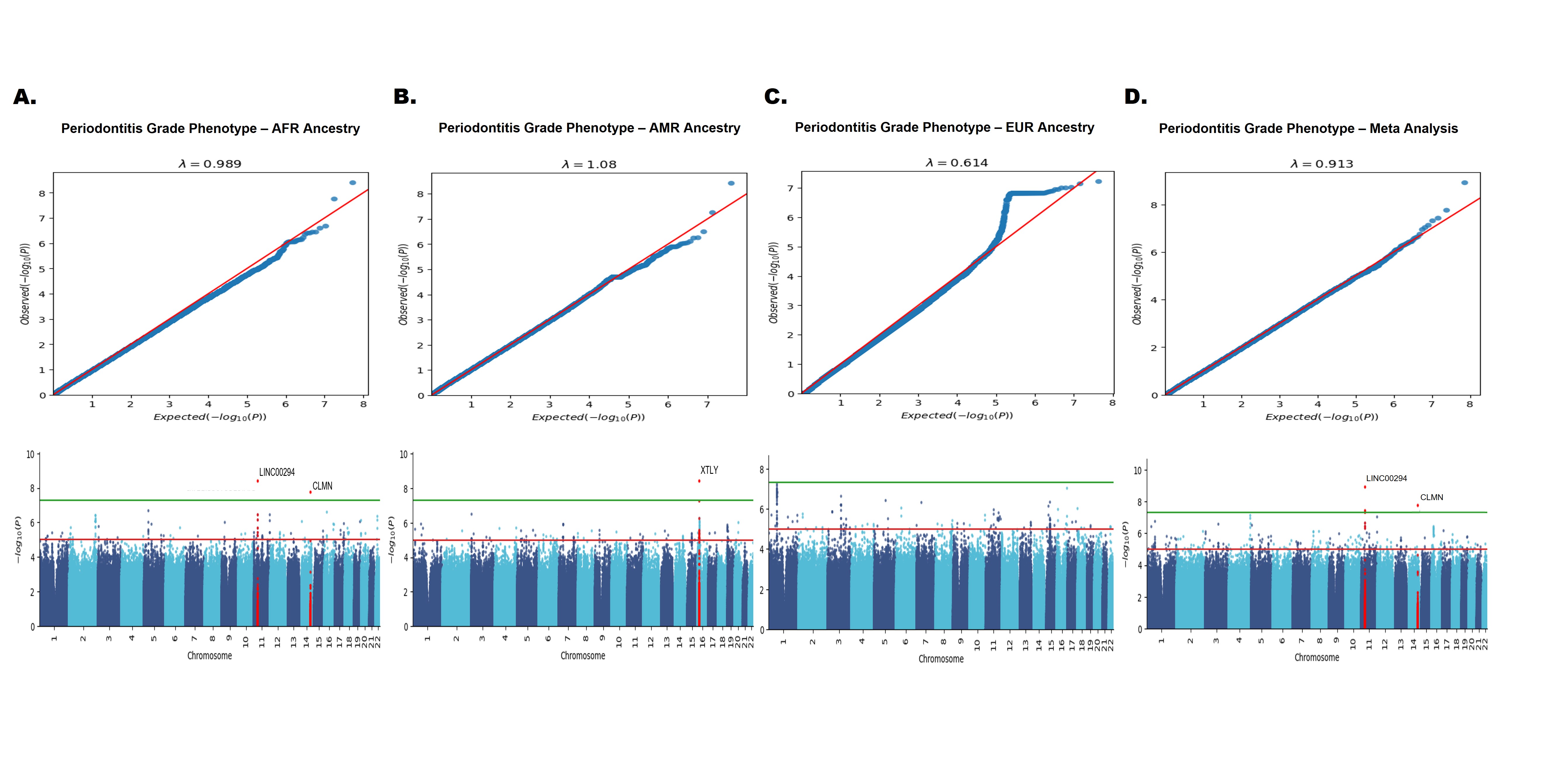

### Supplement Figure 4. Periodontitis Stage Phenotype GWAS Manhattan plots

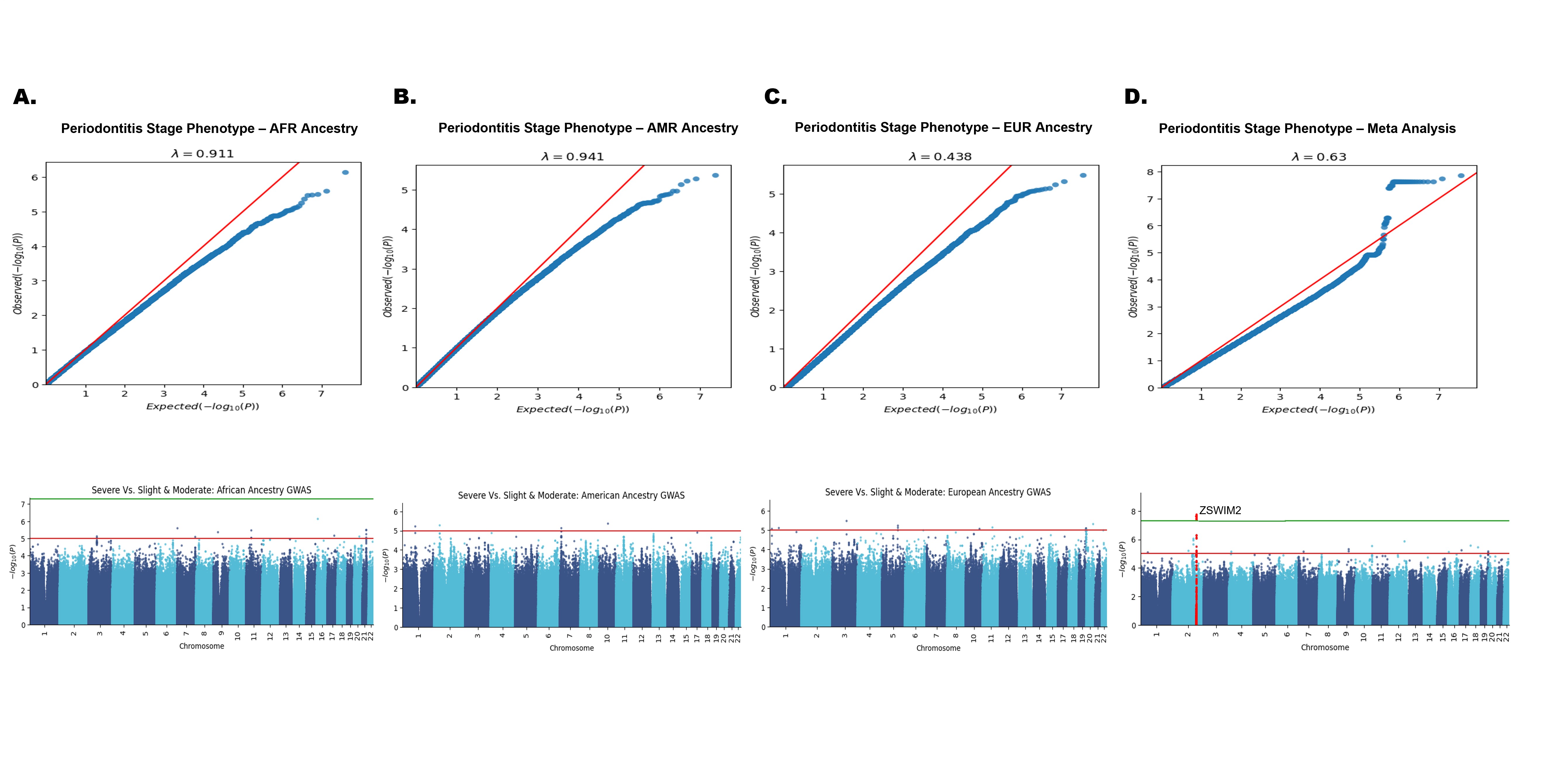

### Supplement Figure 5. UCSC Genome Browser highlighting LINC00294

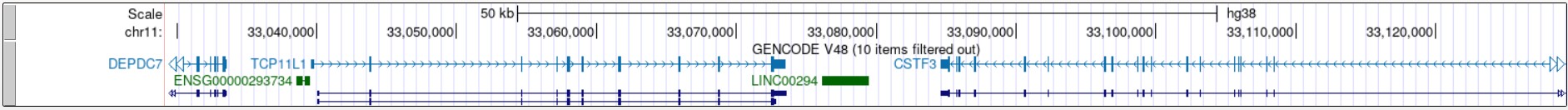
